# Factors predicting Visual Acuity Improvement after Refraction and Reading Ability at 1.5 M in Patients with Low Vision

**DOI:** 10.1101/2025.10.07.25337529

**Authors:** Daniel Martins Cabal, Manuela Molina Ferreira, Rosália Antunes Foschini

## Abstract

**Purpose:** This retrospective study aimed to identify factors predicting improvements in presenting visual acuity (pVA) after refraction and in reading ability among low-vision (LV) patients aged 50 and older.

**Objectives:** To identify predictors of improvement in pVA after refraction and predictors of reading rehabilitation in LV patients.

**Methods:** This observational and retrospective study analyzed medical records of patients aged 50+ years from a LV rehabilitation center in Ribeirão Preto, SP, Brazil, between 2009 and 2019. Improvement in pVA was considered positive if there was an increase of at least 2 logMAR lines after manifest refraction. Reading rehabilitation was deemed successful if the patient was able to read 1.5 M-sized texts.The study evaluated the following predictors: age, sex, pVA, best-corrected visual acuity for distance (dBCVA), the anatomical region responsible for LV, educational level, spherical equivalent, and the degree of cylinder. Data analysis and associations were determined using logistic regression, with a p-value of 0.05 set as the threshold for statistical significance.

**Results:** There were 558 patients with a mean age of 67 years; predominant retinal involvement; the pVA improved after refraction in 20.3%; pVA and the spherical equivalent are predictors of improving pVA after refraction. Among 346 literate patients, 253 (73.1%) could read 1.5 M texts after rehabilitation. The predictors of reading rehabilitation were dBCVA, anatomic region retina, better educational level, and female sex. The worse the dBCVA and the older age hinder reading rehabilitation. Conclusions: The predictors of improvement in pVA after refraction and reading rehabilitation in LV patients highlight the importance of clinical and demographic factors. While age, sex, and education level were not associated with improvements after refraction, both pVA and spherical equivalent emerged as significant predictors. A higher educational level, female sex, and retinal involvement were associated with a greater likelihood of successful reading rehabilitation. Conversely, increasing age and a decrease in dBCVA were linked to a reduced likelihood of reading rehabilitation. These findings underscore the importance of addressing clinical, demographic, and socioeconomic factors in the rehabilitation of individuals with LV.

## INTRODUCTION

Visual impairment increases with age.(1-4) In developing countries, approximately 82% of blind people and 65% of people with mild to severe visual loss were 50 years or older.(5) In 2017, the International Agency for the Prevention of Blindness reported that in Brazil (Tropical Latin America group comprising Brazil and Paraguay) there are 719,140 people with blindness and 3,487,324 people with moderate to severe visual impairment. Visual impairment, particularly in its moderate, severe, or worse forms significantly impacts quality of life across various domains.(6) Globally, it is estimated that approximately 293 million people have moderate or severe visual impairment (presenting visual acuity for distance (pVA) of 20/60 or worse in the better-seeing eye, or visual field of less than 20º, and 43.3 million people worldwide are blind.(1)

According to the 2019 update of ICD-10, moderate visual impairment is defined as presenting visual acuity (pVA) in the better-seeing eye of less than 0.3 and greater or equal to 0.05, or visual field less than 20°.Under ICD-11, moderate visual impairment is pVA in the better-seeing eye of less than 0.5 and greater than or equal to 0.05 or visual field less than 20°. Blindness is defined as pVA less than 0.05, or visual field less than 10°in both ICD-10 and ICD-11. (7) Rehabilitation services for people with sslow vision (LV) or blindness—conditions corresponding to moderate and severe visual impairment and blindness under ICD-10^—^are essential, yet persistent barriers remain. Despite global initiatives, including in Brazil, access is uneven, and the availability of specialized professionals is limited.(8) Addressing these challenges is crucial to mitigate the negative impacts of visual impairment. In the assessssment of visual function, the ophthalmologist may attempt to the opportunity to prescribe glasses or optical aids that may optimize their residual vision,(9) which may improve independence in performing stasks, and promote reintegration into various social spheres. (10, 11) The refraction exam is essential to try to improve residual visual function, with the patient using or not previous optical correction. Fonda described in 1987 and 1994 that 15% of LV patients and 29 % of patients with diabetic retinopathy improved distant VA through refractometry.(12, 13) Sunness et al.(14) described an improvement of greater than 2 logMAR lines in 11% of LV patients in 739 patients with a median VA of 20/80^-2^; and Guo(15) reported an improvement of 27.9%, however, mild LV patients were included. Nevertheless, there is still little data on this subject in developing countries, where socio-economic status and low educational level may interfere in the results of the rehabilitation approach.(16)

There is still little data on improvements in distance VA after refraction, especially after the advent of new therapies for glaucoma and retinal diseases, such as minimally invasive procedures and drugs that reduce abnormal blood-vessel growth. (17, 18) Moreover, few studies report reading-rehabilitation outcomes for LV populations as a whole. Specific groups are commonly evaluated, such as the ones with age-related macular degeneration,(19) but they do not reflect the reading ability data in a general visual rehabilitation service, with many ophthalmological or systemic conditions culminating in LV.(20-22) According to Stelmack et al.,(23) many activities of daily living are closely tied to reading tasks, including checking emails, verifying grocery prices, reading recipes, reading small or fine print, interpreting a thermometer or looking at a menu, and reading handwritten material. The U.S. ISBN Agency (24) further notes that, for most printed books, body text typically uses a font size between 10 and 12 points. According to the National Academies, (25) 1.0 M corresponds to ∼8-point type with an x-height ≈ 1.45 mm; 1.25 M corresponds to ∼10-point; and 1.6 M to ∼12.5-point. In this context, this study aims to determine: 1-the proportion of LV patients who improved their pVA after a manifest refraction for distance vision, and predictors associated with this improvement; 2-the proportion of LV patients who, after rehabilitation, could read text at or below 1.5 M—a threshold generally sufficient for many reading-related activities of daily living and for standard 12-point book print - and the associated predictors.

## MATERIALS AND METHODS

This was a retrospective observational, and analytical study, focusing on patients 50 years or older who were assisted for LV at the Rehabilitation Center (CER) of the Hospital das Clínicas, School of Medicine of Ribeirão Preto – USP (HCFMRP), from 2009 to 2019. The data were accessed from September 1, 2019, to January 1, 2024. It was approved by the Ethics Committee of Clinical Hospital at the Ribeirão Preto Medical School, University of São Paulo (number 58577316.8.0000.5440), which waived the informed consent due to its retrospective nature and followed the tenets of the Declaration of Helsinki. The prerequisites for patients to be referred to this LV service at that period were that they had a pVA of LogMAR > 0.5 or visual field defects leading to visual impairment. Approval was granted by the Research Ethics Committee of HCFMRP, number 5,409,964. The application of free and informed consent form was waived due to its retrospective nature.

The demographic data collected included age, sex, educational level (divided into three groups: illiterate or semi-illiterate, up to incomplete elementary school; and complete elementary school or higher – educational level was informed by the patient during the ophthalmologic exam, or during their registry in the hospital). Clinical data included pVA (VA without glasses for distance or with the current glasses, in both eyes); dBCVA (obtained after the refraction in both eyes, whenever possible: in our LV Sector, dBCVA is routinely measured using logMAR or decimal VA notation charts, and the VA considered is the one which the patient can read half or more of the optotypes on the same line. The decimal VA notation was later transformed to logMAR. When VA was described as hand movements, it was classified as logMAR 1.9); anatomical region that led to LV in the better seeing eye (divided in the following regions according to Gilbert: (26) all the globe, cornea, lens, uvea, retina, optic nerve, others, and globe with normal appearance); refraction data (spherical equivalent, which is the spherical power added to half cylindrical power, and the cylindrical power solely), and near visual acuity (nVA) (categorized into inability or ability to read texts sized less than or equal to 1.5 M). Reading ability was considered positive if there was no indication of difficulties such as “reading with difficulty or very slowly,” “only recognizing numbers,” “spelling separate letters” or “reading separate words but not phrases.” Standardized texts in Portuguese (27-29) were previously printed in Times New Roman font, in sizes 10 to 24 points. The printed 12-point texts were considered as 1.5 M. MNREAD-P reading chart (Minnesota Low Vision Reading Test – Portuguese)(30) was also employed in the rehabilitation process, although without the aim of looking for critical print size or reading speed. In MNREAD-P chart, if patients could read only 1.6M sentences and not smaller ones, they were considered as having no ability to read texts sized less than or equal to 1.5 M. The near reading distance was usually chosen by the patient during rehabilitation, the one providing the best image sharpness, with the use of the glasses or optical aids for near vision. The printed 12-point texts were considered as 1.5 M, therefore, being able to read texts sized 1.6 M or more were considered incapable of reading 1.5 M sized texts. For logistic regressions analysis, spheric equivalent and cylindrical power were annotated in module. The prescription of glasses for distance was also annotated (yes or no).

Eligibility criteria included pVA in the better-seeing eye worse than 0.5 logMAR (≈ decimal acuity < 0.32) but better than light perception, provided the cause of the reduced visual acuity was not treatable, together with a complete ophthalmologic examination performed by an ophthalmologist. Conversely, exclusion criteria encompassed patients with VA limited to light perception or with no light perception, patients with treatable causes of low vision, those with incomplete ophthalmologic evaluation or without examination by one ophthalmologist.

Two outcomes were analyzed: 1) Distant VA improvement greater than or equal to 2 lines in the pVA of the eye with the better dBCVA after manifest refraction (for this analysis, distant pVA was compared with dBCVA obtained after refraction). For this outcome, the following predictors included age, sex, educational level (divided into 3 levels), pVA, spherical equivalent, cylindrical power and anatomical region; 2) The ability to read texts sized smaller than or equal to 1.5 M. Improvement in near VA was calculated by comparing near pVA (using current correction for near) with the post-rehabilitation near VA achieved using best refraction for distance combined with plus lenses and/or optical aids. For this outcome, the following predictors included age, sex, educational level (divided into 2 levels - illiterate and semi-illiterate patients were excluded in this analysis), dBCVA, spherical equivalent, cylindrical power, and anatomical region. Logistic regression was used to analyze predictors (Stata (Stata/IC 15.1, College Station, TX), with significance if p < 0.05. Data were presented as p-value, odds ratio (OR) and 95% confidence interval (95%CI). For the multivariable analysis, only variables with a p-value < 0.20 in the univariable analysis were included. We used the Spearman test to assess the correlation between pVA or dBCVA and age. For the statistical analysis, the eyes with the best dBCVA of each patient and their respective refraction errors were used. If dBCVA was the same bilaterally, the eye with better pVA was selected. When both pVA and dBCVA were identical, we used refractive error from the right eye.

## RESULTS

Among 637 patients attended at CER, HCFMRP, USP, between 2009 and 2019, 46 were excluded due to incomplete data, and 33 due to blindness or light perception. A total of 558 LV patients (87.6% of our total sample), from 50 to 95 years (67.8 ± 11.0 years) were included. There were no differences between sexes (p 0.53 chi-square test). A total of 111 out of 546 (20.3%) improved pVA greater than or equal to 2 logMAR lines for distance after manisfest refraction; and 253 out of 346 (73.1%) read texts sized less than or equal to 1.5 M. There was no correlation between pVA or dBCVA and age (p 0.109 and p 0.06, Spearman test). Age showed no association with pVA, even with their stratification into age groups 50–59, 60–69, 70–79, and 80 years or older (p = 0.964, Fisher’s exact test). Of the 558 patients, 483 (86.6%) had educational level data, with 78 (16.2%) classified as illiterate or semi-illiterate, 290 (60.0%) with incomplete elementary education and 115 (23.8%) with complete elementary education or more. Table 1 shows the study sample demographic and clinical data.

**Table 1:**
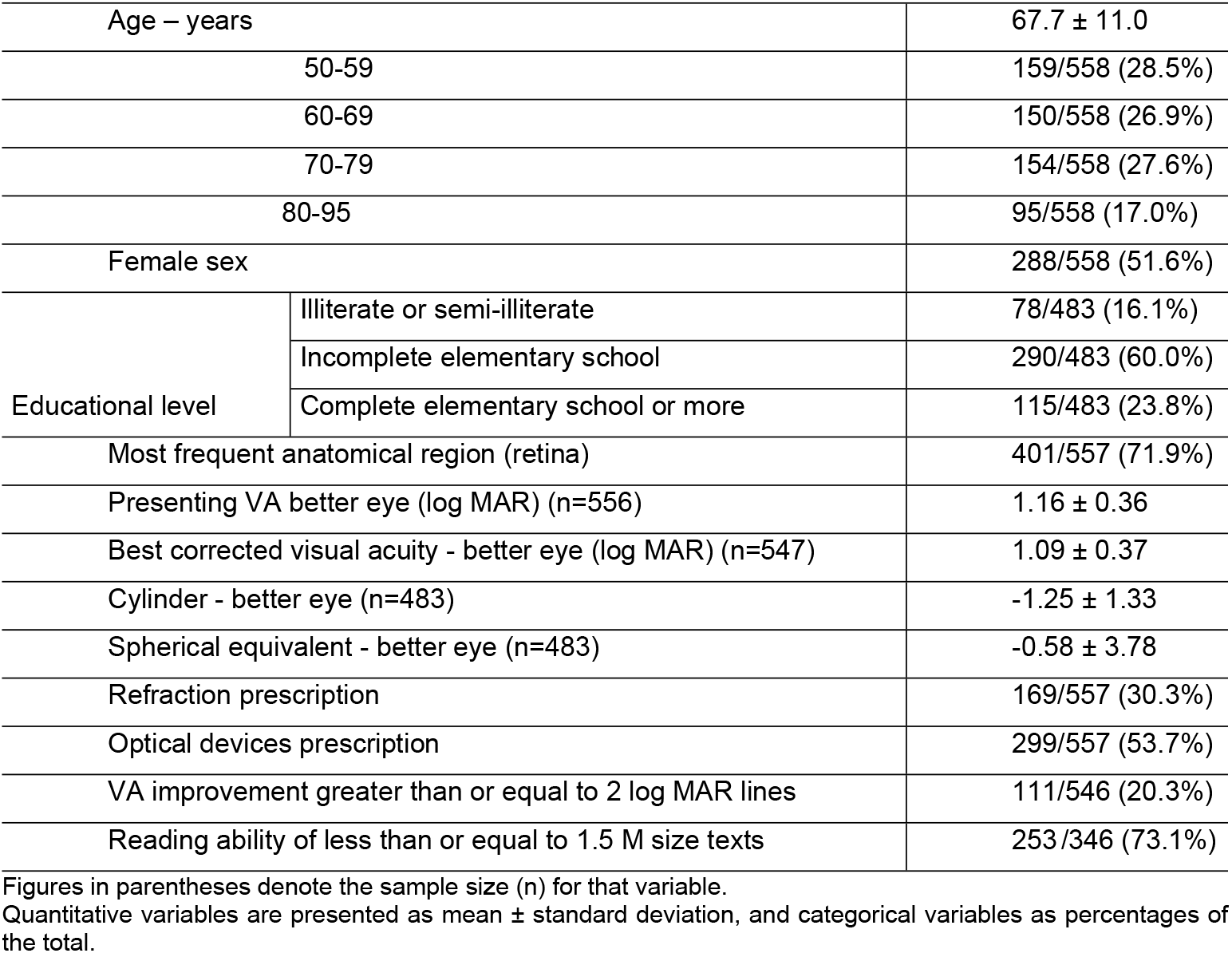
Clinical and demographic data of study patients (n=558)

Table 2 presents the univariate and multivariate logistic regression analyses related to the presenting distant visual acuity improvement after manifest refraction.

**Table 2:** Predictors of presenting distant visual acuity improvement after manisfest refraction.

| Variable | Univariate analysis |  |  | Multivariate analysis<br>(n=476) |  |  |
| --- | --- | --- | --- | --- | --- | --- |
|  | OR | IC 95% | p | OR | 95% IC | p |
| **pVA\* (n=546)** | 1.06 | 1.00 – 1.12 | 0,051 | **1.13** | **1.06 – 1.21** | **0.000** |
| **pVA\*\*profound LV (n=122/546)** | **1.00** | **Reference** |  | --- | --- | --- |
| **pVA\*\* severe LV (n=140/546)** | **4.62** | **1.84 – 11.61** | **0.001** | --- | --- | --- |
| **pVA\*\*moderate LV (n=284/546)** | **7.32** | **3.10– 17.32** | **0.000** | --- | --- | --- |
| Male sex (n=546) | 1.12 | 0.74 – 1.70 | 0.590 | --- | --- | --- |
| Age (n=546) | 1.00 | 0.98 – 1.02 | 0.953 | --- | --- | --- |
| E.L. (illiterate) (76/476) | 1.00 | Reference |  | --- | --- | --- |
| E.L. (incomplete elementary) (286/476) | 1.60 | 0.79 -3.22 | 0.186 | --- | --- | --- |
| E.L. (complete elementary to superior level) (114/476) | 1.49 | 0.68 -3.28 | 0.317 | --- | --- | --- |
| **Sph. Eq.\* (n=478)** | **1.08** | **1.01 -1.15** | **0.018** | **1.07** | **1.00-1.14** | **0.035** |
| **Cylindrical power# (n=478)** | **1.17** | **1.00 – 1.36** | **0.045** | --- | --- | --- |
| Anatomic region (all the globe) € (n=56/543) | 1.00 | Reference |  |  |  |  |
| Anatomic region (cornea) (n=11/543) | 4.18 | 1.18-14.80 | 0.138 | 3.92 | 0.72-21.21 | 0.113 |

\*Increases of 0.1logMAR units; EL: Education Level; LV: low vision; IE: Incomplete Elementary; Esc.; CE: Complete Elementary; Sph. Eq: Spherical equivalent; # Spheric equivalent and cylindrical power in module; € Reference: Glaucoma/all the globe
\*\*pVA: pVA also evaluated based on the categories of visual impairment: profound (logMAR 1.4-1.9); severe (logMAR 1.1-1.3); moderate (logMAR ≤1.0).

Regarding the reading ability, 253 (73.1%) out of the 346 patients read texts sized less than or equal to 1.5 M, and 208 (60.1%) read texts sized less than or equal to 1.0 M. Regarding the 253 patients who read 1.5 M texts, 65 (25.7%) could read only 3 to 16 M sized texts before reading rehabilitation, 100 (38.9%) could read 1.6 to 2.5 M, and 96 (37.9%) already read texts of 1.5 M or smaller, but would like to improve their visual functionality. Table 6 shows the predictors of reading rehabilitation.

**Table 6:**
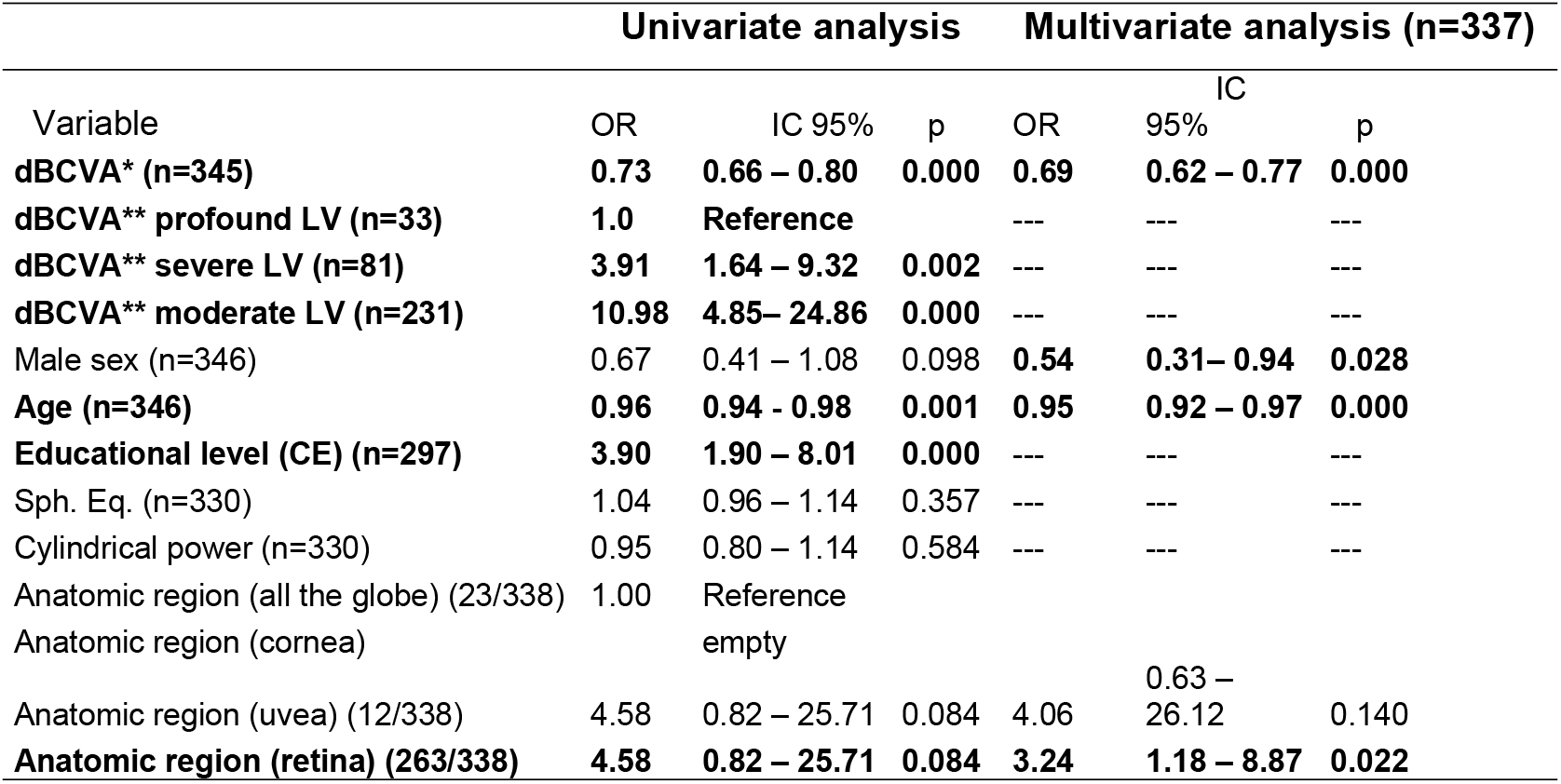
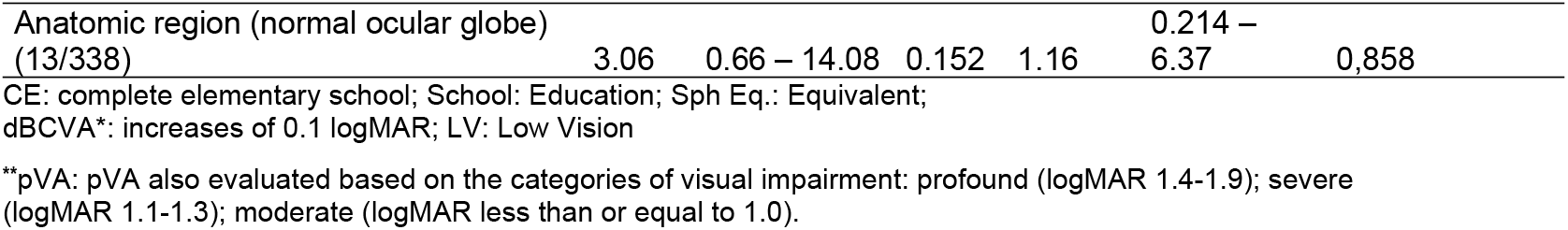
Predictors of reading rehabilitation for texts less than or equal to 1.5M in size.

## DISCUSSION

Global data on LV and rehabilitation are still unsatisfactory, with an even greater gap in developing countries. Data from the Leiden study (3) in people aged 85 and over indicate that visual impairment predicts accelerated physical deterioration in the elderly. Current data show a 34% increase in the risk of elderly people with visual impairment having a low self-assessment of their mental health and a 22% increase in symptoms of loneliness.(31) Another study showed a 47% increase in the risk of dementia in visually impaired elderly people.(32) Thus, understanding the various facets of rehabilitation for LV people may improve visual functionality and physical and cognitive spheres, enhance quality of life, and reduce mortality.(3)

In addition to benefiting the population, rehabilitation centers also provide data on their comorbidities and social and educational data, which allow future decision-making, including better planning to serve the visually impaired. Regarding Brazil, a country with continental dimensions, regional studies can contribute even more, clarifying aspects of the population’s needs and demands. (33-35) Contrary to population studies showing that the mean VA decreases with aging, (36)we found no correlation between pVA or dBCVA and age.

In our sample, 111 (20.3%) patients improved distant pVA greater than or equal to 2 logMAR lines. In a study of 707 old patients, (37) 38.8% did not have their refractive errors corrected, and the non-use of corrective optical lenses was even more frequent in those with ocular disease. In a case series of patients with LV of various etiologies and a mean logMAR VA of 0.92 ± 0.37, the optical correction with glasses improved VA in 33.8% of patients. (38) Sunness and Annan(14) reported an improvement in VA greater than or equal to 2 logMAR lines after refraction in 11% of 739 patients with VA of 20/80^-2^ (or logMAR 0.6). Guo et al.(15) reported improvement of greater than or equal to 2 logMAR lines in 27.9%; however, patients with VA < 0.5 (or logMAR > 0.3) were included (patients with mild LV were included). This data reinforces the need to perform refractometry in the population with LV since there is evidence that improving visual function through optical correction can also reduce symptoms of depression. (39) Regarding quality of life, the data are conflicting. Studies with LV patients show positive results. (40) Still, a Cochrane review on quality of life, conducted in adults with visual impairment, mainly in advanced age and living in high-income countries, found only low and moderate certainty evidence, respectively, of a small benefit in the quality of life related to the rehabilitation of these patients. (5)

Although several studies, including systematic reviews, show a higher proportion of women with LV than men,(1) ours shows no difference in the proportion of LV between sexes, and no relationship between pVA improvement and sex after refraction. However, our study differs from prior work in that it is not population-based; rather, it represents a cohort of patients seen at a low-vision referral center. Also, in our sample, the level of education does not predict changes in pVA after refraction. Therefore, even LV illiterate people should be encouraged to take a refraction exam. The population that improved their distant pVA with refraction had a median spherical equivalent, in module, of 1.87 (interquartiles 1.00 to 3.75 D), similar to those observed in the Brazilian population (hyperopia of 1.61±1.27 and myopia of 1.93±2.38).(41) For every 1.0 D increase in spherical equivalent (regardless of hyperopic or myopic correction), the likelihood of improving pVA by 0.1 logMAR increases by 8% (OR = 1.08, 95% CI: 1.00–1.15) in univariate analysis and by 7% (OR = 1.07, 95% CI: 1.00– 1.14) in multivariate analysis. Guo et al.(15) reported similar findings: for every 1.0 D increase in spherical equivalent, the likelihood of improving VA by ≥2 logMAR lines increased by 5% (OR = 1.05, 95% CI: 1.01–1.09). Regarding cylindrical power, our analysis showed that each 1.0 D increase was associated with a 17% higher chance of improving pVA by ≥2 logMAR lines (OR = 1.17, 95% CI: 1.00–1.36). Although many LV patients do not perceive significant improvement in visual resolution with optical correction, the potential for reducing blurring underscores the importance of performing a refractive exam.(42)

In our study, most LV patients had the retinal involvement (71.9%). Similarly, Leasher et al, (43) in a population-based study, identified macular degeneration and glaucoma as the leading causes of irreversible LV in South America. Data from the World Health Organization from 2023 (44) also highlight retinal diseases, such as diabetic retinopathy and macular degeneration, as major contributors to LV. We found no association between distant pVA improvement after refraction and the affected anatomical region. However, a comparable study involving 2,923 participants reported an association with corneal diseases, showing that 34.8% of patients with corneal conditions experienced VA improvement with refraction. (15) Our study may have lacked sufficient power to detect a similar association with corneal diseases.

Regarding reading ability, after rehabilitation, 253 patients (73.1%) were able to read texts less than or equal to 1.5 M again. A study on reading rehabilitation reported an even higher proportion, with 94% of patients regaining the ability to read 10-point font texts; however, their average VA was 0.18 ± 0.15 (decimal scale; approximately 0.7/0.8 in logMAR VA scale), and all had age-related macular degeneration. (19) Another study found that only 2% of LV patients could read a newspaper without optical aids, whereas with optical aids, approximately 51% were able to do so. (45)

We observed a direct association between dBCVA and reading rehabilitation. Higher dBCVA was linked to an increased likelihood of reading less than or equal to

1.5 M. Additionally, dBCVA may also influence reading speed. (19, 46) However, dBCVA is not the sole predictor of reading rehabilitation. Other visual function factors, such as contrast sensitivity, scotoma size, and visual field (number of letters perceived simultaneously), are also known to impact rehabilitation outcomes.(47) In our study, women demonstrated a higher likelihood of successful reading rehabilitation than men (Table 6). In a study with individuals with normal vision and logMAR VA less than or equal to 0.1,(48) fifteen measures of visual perception were evaluated, including VA; men significantly outperformed women in six of these tests, while women did not outperform men in any. This suggests that additional factors may be influencing our findings. Cultural influences could play a role, as women often engage in activities that require good near vision, such as grocery shopping - where they need to check prices, expiration dates, and product ingredients - or managing family members’ medications. Additionally, a survey found that although women make up 51% of Brazil’s population, they account for approximately 57% of book buyers.(49)

We found that with each additional year of age, the likelihood of reading texts less than or equal to 1.5 M decreases by 4%. Other studies also confirm that reading ability and speed decline with aging.(50, 51)

Our study may have lacked sufficient power to detect an association between reading rehabilitation and certain anatomical regions, such as the cornea. Additionally, neither spherical equivalent nor cylindrical power appeared to be linked to reading rehabilitation. However, we did not evaluate whether the astigmatism with-the-rule or against-the-rule could influence rehabilitation outcomes. Notably, one study indicated that with-the-rule astigmatism tends to have a greater negative impact on VA and near vision than against-the-rule astigmatism.(52)

Regarding the anatomical region, retina is the most frequent affected area in LV patients. Among those who were able to read, 195 (77.7%) had retinal involvement (Tables 1 and 6). Our findings align with a Cochrane systematic review, (5) which identified the retina - particularly due to age-related macular degeneration as the most common cause of LV in the elderly. The present study may not have had adequate power to detect an association between reading rehabilitation and cornea. Previous research has demonstrated a significant improvement in reading speed among patients with corneal diseases when using optical resources. (53)

The limitations of this study stem from its retrospective nature. Notably, educational background data was missing in approximately 15% of medical records, and there was no information on whether patients obtained the prescribed distance glasses or if they consistently used optical aids for reading. However, a key strength of this study is the standardized rehabilitation process, as it was conducted by only two ophthalmologists and one orthoptist throughout the study period.

In conclusion, 20.3% of patients showed distant pVA improvement after manifest refraction; among literate patients, 73.1% were able to read texts less than or equal to1.5 M after rehabilitation; in the multivariate analysis, pVA and spherical equivalent were identified as predictors of a 0.2 logMAR lines improvement in pVA after distant refraction; in the multivariate analysis, increasing age and decreasing dBCVA reduced the likelihood of reading rehabilitation, whereas completing elementary school and female sex were associated with a greater chance of regaining reading ability.

## Data Availability

If the manuscript is accepted, we can share data publicly using redcap or some other option offered by the journal

